# Association of Transcoronary Cytokine Gradients with Pericoronary Adipose Tissue Attenuation: A Transcoronary and Transcardiac sampling study

**DOI:** 10.64898/2026.08.06.26359918

**Authors:** Neville Tan, Graeme Lancaster, Frank Du, Shaun Khanna, William Chan, Nitesh Nerlekar, Thomas H. Marwick

## Abstract

**Background:** Pericoronary adipose tissue (PCAT) attenuation on coronary computed tomography angiography (CTCA) has emerged as a novel non-invasive biomarker of coronary inflammation and cardiovascular risk. The degree to which PCAT reflects local or systemic inflammation remains uncertain. We hypothesized that the presence, location and extent of PCAT would be associated with transcoronary or transcardiac cytokine gradient.

**Methods:** This prospective cohort study involved 31 adults with stable coronary artery disease who underwent clinically indicated CTCA within 90 days of invasive coronary angiography. Patients with acute coronary syndromes or unstable angina were excluded. Blood samples were obtained from peripheral vein, coronary sinus, aortic root, and right coronary artery at time of cardiac catheterization. Plasma interleukin-6 (IL-6) and interleukin-1β (IL-1β) concentrations from each site were used to calculate transcardiac and transcoronary cytokine gradients. PCAT attenuation was measured using semi-automatic software by readers blinded to clinical and biochemical endpoints.

**Results:** Participants were predominantly male (76%), aged 66.6 ± 9.4 years, with a high prevalence of hypercholesterolemia (76%), hypertension (73%), and diabetes (36%). Mean PCAT attenuation was −74.8 HU (RCA), −70.3 HU (LCx), and −73.6 HU (LAD). Regression analyses showed no significant associations between PCAT attenuation and IL-6 gradients across any coronary territory (all p >0.40; R² ≈ 0), including in plaque-free subgroup analyses. IL-1β was below the assay detection limit in 81% of participants; analyses using non-parametric testing and logistic showed no association with PCAT attenuation. RCA (OR 0.96, 95% CI 0.88–1.06, p=0.46), LCx (OR 1.00, 95% CI 0.91–1.09, p=0.94), LAD (OR 0.99, 95% CI 0.90–1.08, p=0.81).

**Conclusion:** In a cohort with predominantly stable coronary disease, PCAT attenuation was not associated with intracardiac or intracoronary IL-6 or IL-1β gradients, including in plaque-free vessels. These findings suggest that PCAT attenuation may not reflect active cytokine-mediated coronary inflammation in stable disease.

## Introduction

Peri-coronary adipose tissue (PCAT) has gained increasing recognition as a non-invasive marker of cardiovascular risk^1 2 3^. Coronary computed tomography angiography (CCTA) has enabled non-invasive quantification of PCAT attenuation radiodensity, reflecting alterations in adipocyte size, lipid content.^1,4^ Increased PCAT attenuation has been associated with adverse clinical outcomes including major adverse cardiac events, mortality, and high-risk plaque features^1^. These observations have positioned PCAT as a promising imaging biomarker of coronary inflammation not only in acute coronary syndrome, but also as a risk modifier in early coronary artery disease^4,5^.

As PCAT shares a blood supply with the vasa vasorum of coronary arteries^6 7^, it both responds and also participates in endothelial inflammation and injury.^8^ Adipose tissue produces various bioactive molecules including the inflammatory cytokines TNF-alpha, Il-1B and Il-6, and has a distinct role in regulating cardiometabolic function^9^. The downstream effect of PCAT-derived inflammatory cytokines could contribute towards plaque progression^2^. The proximity of PCAT to arterial smooth muscle cells also provides the potential to influence vessel homeostasis and atherosclerosis. In biopsies of peri-aortic adipose tissue obtained at the time of coronary artery bypass grafting, elevated PCAT has been associated with smaller adipocyte size and reduced lipid content ^7^. In addition, cultured pre-adipocytes exhibited impaired differentiation when exposed to pro-inflammatory cytokines (IL-6, TNF-α, IFN-γ) in response to upregulation with angiotensin II. Circulating inflammatory cytokines (IL-7, IL-15, MCP-1) have been shown to be associated with PCAT attenuation on CTCA ^10^. Albertini et al. further demonstrated links between PCAT, peripheral IL-6/TNF-α, plaque burden - and the influence of systemic modifiers such as chronic stress. Together, these studies strongly support the concept that PCAT reflects inflammatory biology relevant to coronary disease.

Coronary sinus sampling is a validated method of isolating local cardiac production from systemic inflammation,^11^ and therefore provides a physiological approach to assess cardiac-specific inflammatory activity by comparing myocardial vs systemic cytokine concentrations. Previous studies have demonstrated significantly higher TNF-α and IL-6 levels in the coronary sinus compared with the aortic root in patients with unstable angina ^12^, supporting evidence of intracardiac cytokine release in acute coronary syndromes. Previous studies have also shown transcoronary gradients for IL-1β, IL-16 and IL-18 in ACS and these gradients were lowered by colchicine^13^, confirming both the biological relevance and potential for therapeutic target of deriving local inflammatory signals. However, to date, no studies have directly examined the presence of a transcoronary inflammatory cytokine gradient or its relationship with PCAT attenuation in patients with stable coronary artery disease. This represents an important mechanistic gap in our understanding of whether increased PCAT attenuation reflects merely a downstream or instead serves as an originator of active inflammatory signalling within the coronary circulation. Demonstration that concentrations of inflammatory mediators are higher in coronary venous blood than in systemic circulation, would provide stronger biological validation for PCAT as culprit of local vascular inflammation. Accordingly, this study aimed to investigate the relationship between pericoronary adipose tissue and coronary inflammation, and to determine whether evidence exists for a unidirectional or bidirectional atherogenic signal, as assessed by the presence of a transcoronary cytokine gradient.

## Methods

### Patient selection

This prospective cohort study recruited 31 adult patients (≥18 years) from July 2023 to November 2024 - with suspected or stable coronary artery disease who had undergone CTCA within 90 days before clinically indicated invasive coronary angiography. The mean interval between CT scan to angiography was 35 days (IQR 26–53 days). This study was designed as a mechanistic, proof-of-concept investigation exploring the relationship between CT-derived PCAT and directly sampled intracoronary inflammatory biomarkers. As this study was designed as an exploratory investigation exploring the relationship between PCAT and intracoronary inflammatory biomarkers, our sample size was based on previous sampling studies assessing trans-cardiac sampling in a similar range ^14 15 16^. Exclusion criteria included myocardial infarction, acute coronary syndrome, unstable angina or acute chest pain syndromes requiring emergency intervention. Patients with previous percutaneous coronary intervention or coronary artery bypass grafting surgery were also excluded. Patients provided written informed consent for use of CTCA images, peripheral and coronary venous blood sampling at the time of study. This study was approved by Research Ethics Committee – Monash Health (Reference - 86262 RES-22-0000-258A).

### Cytokine measurements

Blood samples were obtained from 4 separate anatomical sites 1) peripheral, 2) coronary sinus 3) aortic root 4) right coronary artery during cardiac catheterization, and performed before heparin administration or any interventional procedure was commenced. Specimens were immediately centrifuged, and separated into plasma aliquots according to a standardized protocol. Aliquots were stored at - 80 °C freezer on-site, until batch analysis. Biomarker analyses of were subsequently performed at a core laboratory by experienced personnel that were blinded to clinical and imaging data, plasma IL-1B and IL-6 concentrations were quantified using previously validated assays.

IL-1β is an upstream pro-inflammatory cytokine produced predominantly by activated macrophages through inflammasome activation within atherosclerotic plaques^17^. It plays a important role in amplifying local inflammatory responses, including endothelial activation, leukocyte recruitment, and plaque destabilisation^18^. IL-6 functions downstream of IL-1β and acts as a cascade cytokine linking local vascular inflammation to systemic inflammatory responses, including hepatic acute-phase reactant production. Elevated IL-6 concentrations have been associated with coronary artery disease burden, plaque vulnerability, and adverse cardiovascular events^19^. Previous studies have shown that upstream IL-1β inhibition reduces cardiovascular events and is accompanied by downstream suppression of IL-6 signalling^20^, and both IL-1β and IL-6 have been validated as coronary inflammatory biomarkers in previous coronary sampling studies ^12 13^.

Peripheral cytokine concentrations primarily reflect systemic inflammatory burden and are influenced by numerous non-cardiac factors, including infection and autoimmune disease. We therefore defined the following primary outcomes:

i) Transcardiac gradient (*coronary sinus minus peripheral concentration*) reflecting the net cardiac contribution above systemic background. A neutral or negative gradient suggests that the heart is not a major contributor to circulating cytokine burden, even if absolute levels are elevated.
ii) Transcoronary gradient (*coronary sinus minus aortic root*) indicates cytokine production in the coronary circulation itself. It is relevant to the PCAT hypothesis in that biologically active PCAT causing perivascular inflammation might be associated with a net release of cytokines from the coronary vasculature.
iii) *Local* coronary inflammatory signal was assessed by comparing cytokine concentrations sampled directly from the RCA against the aortic root.

### CT coronary angiography and PCAT

Coronary plaque volumes were quantified in vessels >2 mm diameter using semi-automated software Medis QAngioCT (Netherlands), with manual correction of vessel and luminal contours and plaque classification. Total plaque burden per patient was derived from summation of individual segments. Plaque components were stratified – into non-calcified plaque included necrotic core (−30 to 75 HU), fibrofatty (76–130 HU), and fibrous tissue (131–350 HU), distinct from dense calcified plaque (>350 HU). PCAT attenuation was quantified by a reader blinded to clinical outcome using semi-automated software (Q Angio Research Edition 2.5, MEDIS, Leiden, Netherlands). PCAT was defined as adipose tissue within a radial distance from the outer coronary wall equal to the vessel diameter, using a standardized attenuation threshold (−190 to −30 HU). Measurements were performed in pre-specified segments surrounding the proximal RCA (10–50 mm from the ostium), as well as corresponding segments of the LCx and LAD.

Given the exploratory and mechanistic focus of this study, we opted to use a PCAT attenuation metric rather than incorporating a more complex algorithm-derived measure such as the Fat Attenuation Index (FAI).^1,4^ While FAI has shown incremental value as a prognostic and risk-stratification tool in recent studies, as our primary objective was to examine biological signal correlated with sampled intracoronary and intracardiac cytokine gradients.

### Statistical analysis

Continuous variables are presented as mean ± standard deviation (SD) or median (interquartile range [IQR]), according to data distribution. Categorical variables are presented as frequencies and percentages. Comparisons between groups were performed using the Student’s *t*-test or Mann–Whitney *U* test for continuous variables, as appropriate. Pearson correlation coefficients were used to evaluate linear associations between PCAT attenuation and IL-6 concentrations or cytokine gradients. Univariable and multivariable linear regression analyses were performed to assess the association between PCAT attenuation and IL-6 concentrations, while logistic regression was used to evaluate the association between PCAT attenuation and detectable IL-1β concentrations. A two-sided *p* value <0.05 was considered statistically significant. Statistical analyses were performed using STATA version 18.0 (StataCorp, College Station, TX, USA) and IBM SPSS Statistics version 24.0 (IBM Corp., Armonk, NY, USA).

## Results

### Patient characteristics

Participants were predominantly male (76%), aged 66.6 ± 9.4 years. Cardiovascular risk factors were common - hypercholesterolaemia (76%), hypertension (73%), diabetes (36%) and overweight (BMI 28.7±6.5 kg/m) - and coronary calcium burden was 768 ± 758. Following angiographic assessment, 6 (19%) underwent percutaneous coronary intervention (PCI) and 8 (25%) were referred for coronary artery bypass grafting (CABG).

Severe coronary stenoses were more commonly identified on CT coronary angiography (CTCA) than on invasive coronary angiography across all vessels (**Table 1**). On CTCA, severe lesions were observed in the left anterior descending artery (LAD) in 25 patients (81%), followed by the right coronary artery (RCA) in 15 patients (48%) and left circumflex artery (LCx) in 10 patients (32%). PCAT attenuation was comparable across vessels (**Table 1**). Interobserver reliability was excellent (ICC = 0.96 95% CI 0.89–0.98), without any evidence of systematic bias (mean difference −0.59, 95% CI −1.55 to 0.38; p = 0.22)

**Table 1.**
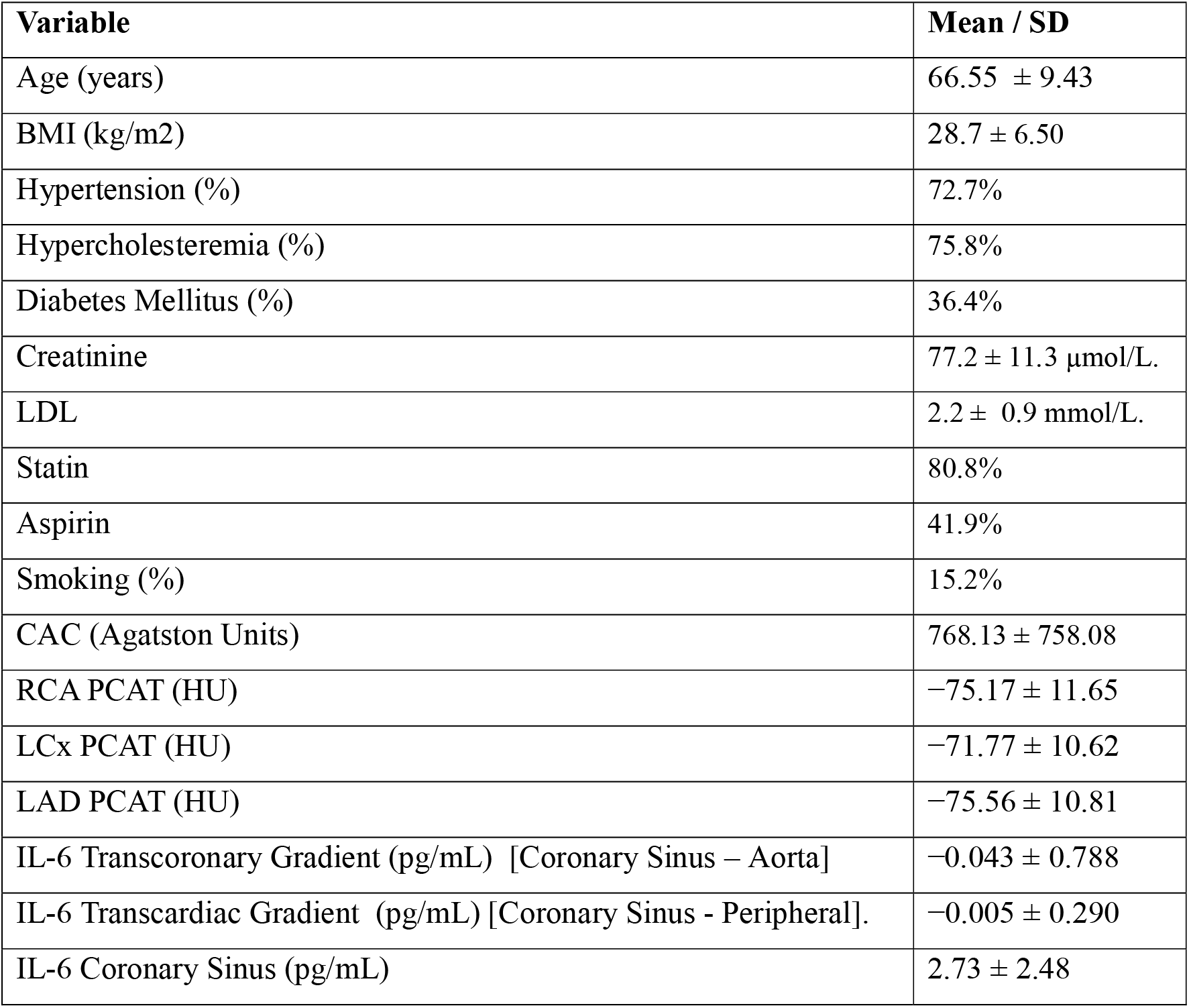
Baseline clinical, biochemical, and imaging characteristics of the study cohort. Continuous variables are presented as mean ± standard deviation, and categorical variables as percentages. IL-6 transcoronary and transcardiac gradients were calculated as (coronary sinus – aortic) and (coronary sinus – peripheral) respectively.

**Table 1B.** Coronary plaque volumes and composition within the study cohort. Continuous variables are presented as mean ± standard deviation.

| Plaque Measure | Mean volume (mm<sup>3</sup>) ± SD |
| --- | --- |
| Total Plaque Volume | 458.53 ± 260.74 |
| Fibrous Plaque | 254.09 ± 122.31 |
| Fibrofatty Plaque | 63.82 ± 42.31 |
| Non-calcified Plaque | 353.20 ± 176.62 |
| Dense Calcified Plaque | 102.27 ± 109.54 |

### Association between PCAT and IL-6

Mean IL-6 transcoronary and transcardiac gradients were small (**Table 1**). Across the cohort, there was no significant association between PCAT attenuation and IL-6 concentration gradients for any coronary territory - RCA, LCx, or LAD. **(Figure 1)**, indicating that PCAT attenuation explained virtually none of the variability in IL-6 gradients. No significant associations were observed between PCAT attenuation and transcoronary IL-6 gradients (coronary sinus – aorta) in any coronary territory (**Figure 2**).

**Figure 1.**
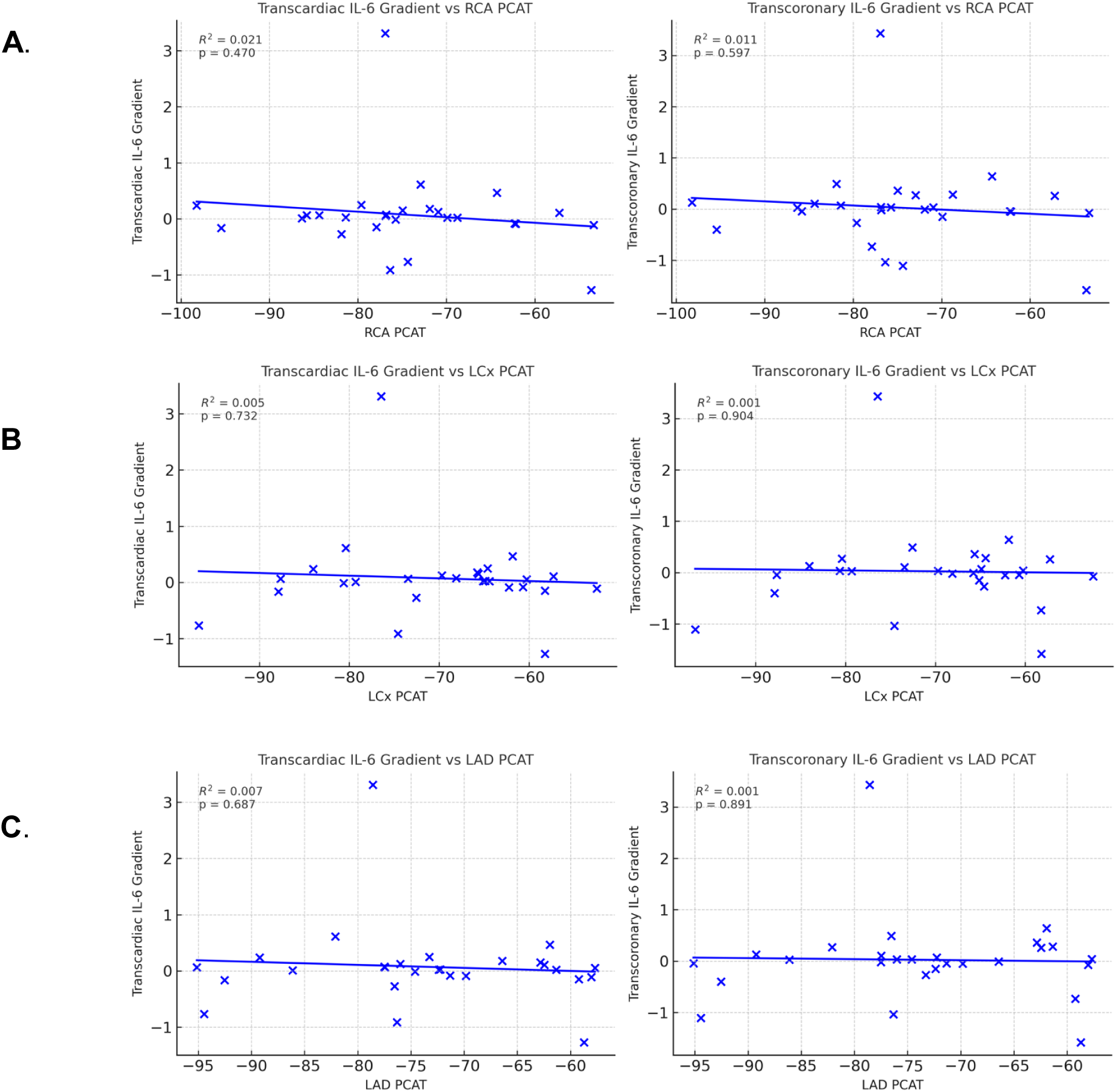
Association between PCAT attenuation and transcardiac and transcoronary IL-6 gradients in the A) Right coronary artery, B) Left circumflex, C) Left anterior descending.

**Figure 2.**
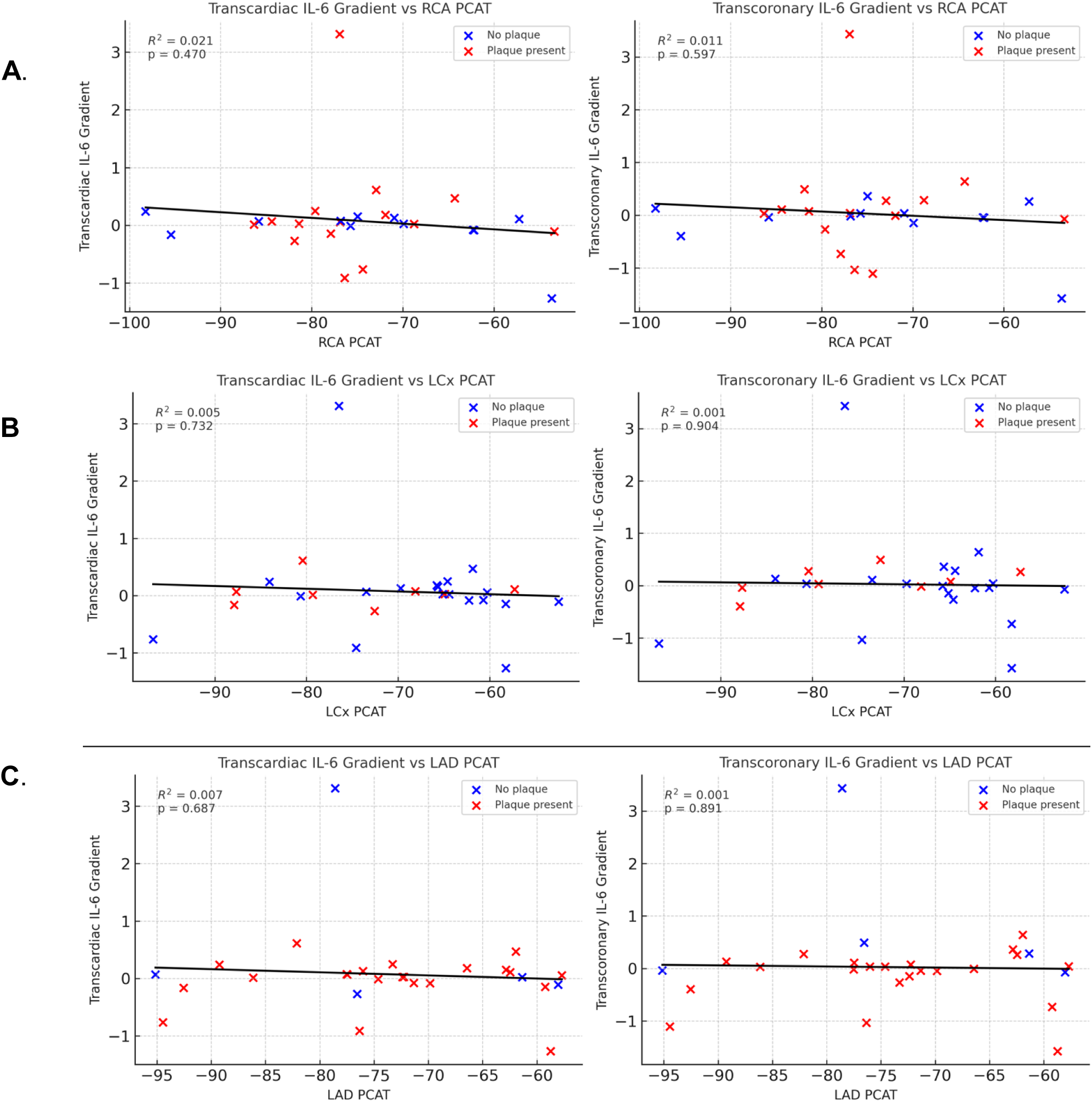
Association between PCAT attenuation and transcardiac and transcoronary IL-6 gradients in vessels with (red) and without plaque (blue) - A) Right coronary artery, B) Left circumflex, C) Left anterior descending.

Finally, we explored RCA PCAT attenuation relates to a *local* coronary inflammatory signal by comparing cytokine concentrations sampled directly from the RCA against the aortic root (RCA – aortic root) (**Figure 3**). This sampling strategy was designed to detect a proximal “outside-in” or perivascular contribution: if PCAT, conventionally measured at the proximal RCA - were metabolically active and releasing cytokines into the adjacent coronary circulation, a measurable step-up relative to the aortic root might be expected, with higher (less negative) PCAT attenuation tracking with a larger RCA–aortic gradient. However, RCA PCAT attenuation did not predict the presence of a local RCA gradient (r=−0.07, p=0.72). The absence of association persisted even when restricting analyses to non-diseased, plaque-free RCA segments to reduce confounding from plaque-driven cytokine production (r=0.08, p=0.785).

**Figure 3.**
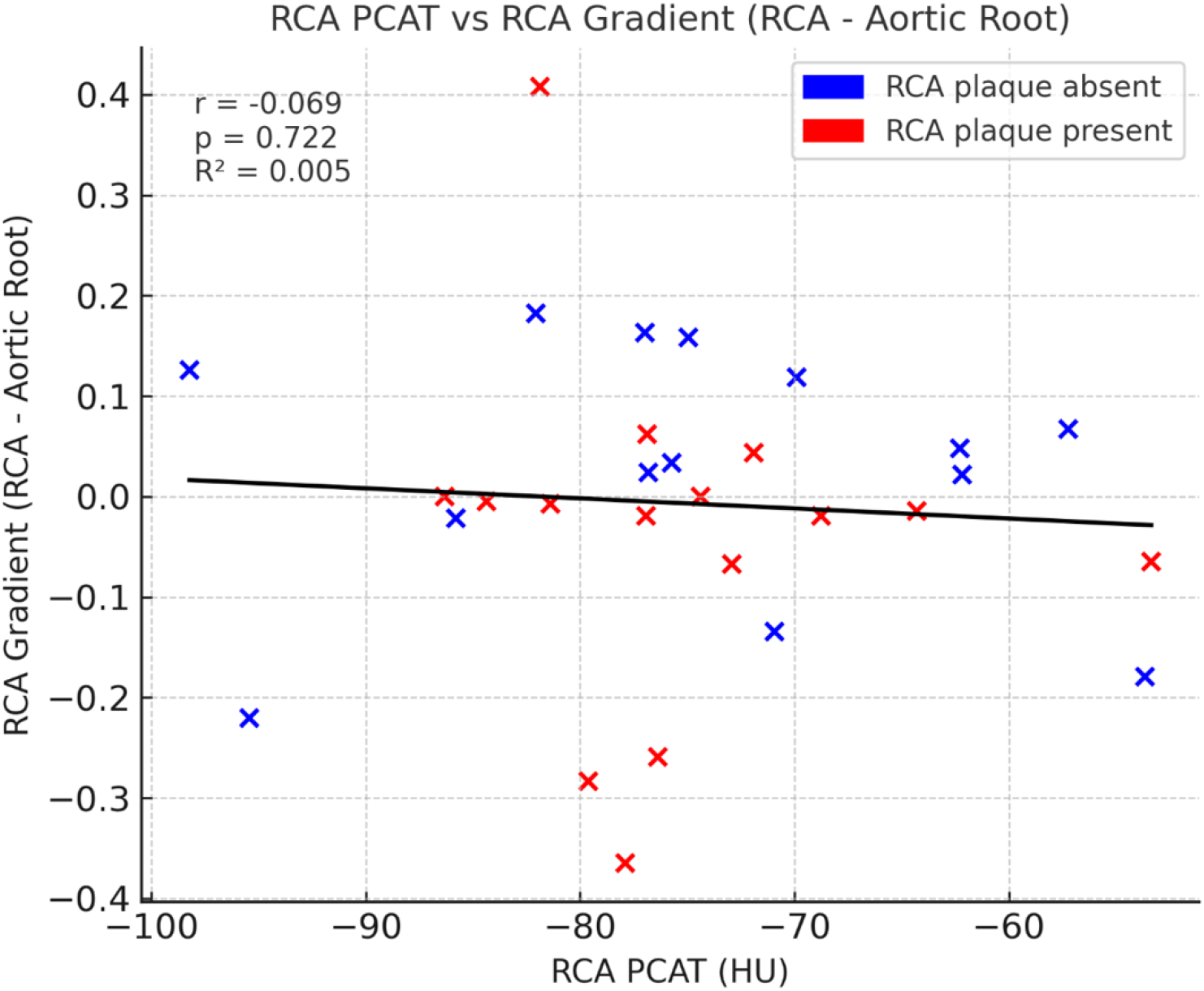
Association between right coronary artery (RCA) pericoronary adipose tissue (PCAT) attenuation and the local coronary inflammatory gradient. (RCA minus aortic root). No significant association was observed in the overall cohort (r = −0.07, p = 0.72), and this remained unchanged when analyses were restricted to plaque-free RCA segments (r = 0.08, p = 0.785), suggesting that proximal RCA PCAT attenuation does not reflect a detectable local intracoronary cytokine signal.

### Association of PCAT and inflammation with plaque features

The mean total plaque volume was 1011.68 ± 508.81 mm³. Among plaque subtypes, fibrous plaque comprised the largest component (563.53 ± 220.78 mm³), followed by dense calcified plaque (224.22 ± 232.84 mm³), fibrofatty plaque (141.66 ± 84.18 mm³), and non-calcified plaque (NCP) (75.92 ± 52.46 mm³).

Regression and Pearson correlation analysis demonstrated no statistically significant associations between vessel-specific pericoronary adipose tissue (PCAT) attenuation and total plaque volume. Correlation coefficients were weak across all vessels (RCA: r = −0.11, p = 0.687; LCx: r = −0.27, p = 0.298; LAD: r = −0.10, p = 0.713). Similarly, no significant associations were observed between PCAT attenuation and fibrous plaque volume in the RCA, LCx, or LAD. All correlations were weak to modest with all *p*-values being above >0.05. (**Table 2**)

**Table 2A.** Association between PCAT attenuation across three major coronary arteries (RCA, LCx, LAD) and coronary sinus IL-6 concentration, Transcoronary Gradient and Transcardiac Gradient. No coronary territory demonstrated statistically significant association with IL-6 (all p > 0.10).

| Predictor | B | SE | Standardized $\beta$ | t | P-value | 95% CI (Lower–Upper) |
| --- | --- | --- | --- | --- | --- | --- |
| **Constant** | −0.97 | 3.99 | — | −0.24 | 0.810 | −9.31 to 7.36 |
| **RCA PCAT attenuation (HU)** | 0.094 | 0.068 | 0.400 | 1.39 | 0.179 | −0.047 to 0.235 |
| **LCx PCAT attenuation (HU)** | −0.210 | 0.125 | −0.813 | −1.68 | 0.108 | −0.470 to 0.050 |
| **LAD PCAT attenuation (HU)** | 0.055 | 0.128 | 0.218 | 0.43 | 0.671 | −0.212 to 0.323 |
| **Transcoronary Gradient** |  |  |  |  |  |  |
| **RCA PCAT** | 0.008 | 0.015 | 0.107 | 0.536 | 0.597 | −0.023 to 0.039 |
| **LCx PCAT** | 0.002 | 0.015 | 0.023 | 0.117 | 0.908 | −0.030 to 0.033 |
| **LAD PCAT** | 0.002 | 0.015 | 0.027 | 0.134 | 0.894 | −0.029 to 0.033 |
| **Transcardiac Gradient** |  |  |  |  |  |  |
| **RCA PCAT** | −0.001 | 0.005 | −0.029 | −0.149 | 0.882 | −0.012 to 0.010 |
| **LCx PCAT** | −0.003 | 0.005 | −0.125 | −0.657 | 0.517 | −0.014 to 0.007 |
| **LAD PCAT** | −0.003 | 0.005 | −0.112 | −0.584 | 0.564 | −0.014 to 0.008 |

**Table 2B.** Correlation of PCAT Attenuation with plaque subtypes (Fibrous, Fibrofatty, Non-Calcified, and Dense Calcified Plaque Volumes)

| Plaque volume | RCA, r (p) | LCx, r (p) | LAD, r (p) |
| --- | --- | --- | --- |
| Fibrous | 0.13 (0.616) | −0.04 (0.865) | 0.13 (0.621) |
| Fibrofatty | −0.18 (0.497) | −0.25 (0.333) | −0.14 (0.599) |
| Non-calcified | −0.17 (0.520) | −0.25 (0.330) | −0.14 (0.592) |
| Dense calcified | −0.24 (0.347) | −0.39 (0.124) | −0.24 (0.345) |

*Values are Pearson correlation coefficients (r), with corresponding p-values*

### Association between PCAT and IL-6

In multivariable linear regression, PCAT attenuation in the RCA, LCx, and LAD was not significantly associated with coronary sinus IL-6 concentration, the transcoronary IL-6 gradient, or the transcardiac IL-6 gradient. For coronary sinus IL-6, RCA PCAT showed a positive but non-significant association (B = 0.094, p = 0.179), LCx PCAT showed an inverse non-significant association (B = −0.210, p = 0.108), and LAD PCAT showed no significant association (B = 0.055, p = 0.671). Similarly, no regional PCAT measure was associated with the transcoronary gradient (all p ≥ 0.10) or transcardiac gradient (all p ≥ 0.10), with all 95% confidence intervals crossing zero (**Table 2**) Since PCAT is measured at the proximal vessel segments – and to assess whether the presence of atherosclerotic plaque might confound the detection of an inflammatory signal, we repeated regression analyses after repeated restricting the cohort to plaque-free vessels only.

In this plaque-free subset of 14/31 patients, associations between PCAT and cytokine gradients remained weak and non-significant for all vessels. For the RCA a modest numerical increase in explanatory power was observed for the transcardiac gradient (R²=0.22), but this did not reach statistical significance (p=0.13) and the association with the transcoronary gradient was also non-significant (R²=0.09, p=0.34). No significant association was observed in the plaque-free subset of LCx and LAD, LCx - (transcardiac: R²=0.008, p=0.71; transcoronary: R²=0.005, p=0.78). LAD, (transcardiac: R²=0.04, p=0.75; transcoronary: R²=0.03, p=0.80) respectively.

### Association between PCAT and IL-1B

Interleukin-1β (IL-1β) concentrations were mostly below the lower limit of detection of the assay; 26 of 31 participants (83%) demonstrated undetectable values. As calculation of transcoronary or transcardiac IL-1β gradients was not feasible, we therefore analysed IL-1β as a binary outcome (detectable vs undetectable) and assessed whether PCAT attenuation predicted detectable IL-1β. There were no significant differences in PCAT attenuation between participants with detectable versus undetectable IL-1β for the RCA (U=78.0, p=0.323), LCx (U=63.0, p=0.889), or LAD (U=67.0, p=0.716). **(Table 3).** Similarly, logistic regression analysis demonstrated no association between PCAT attenuation and detectable IL-1β - Logistic regression demonstrated no significant association for attenuation measured in the RCA (OR 0.96, 95% CI 0.88–1.06, p=0.456), LCx (OR 1.00, 95% CI 0.91–1.09, p=0.941), or LAD (OR 0.99, 95% CI 0.90–1.08, p=0.805) **(Table 3).** Sensitivity analysis demonstrated that the interval between CT and coronary angiography was not associated with either the IL-6 transcoronary gradient (=0.070) or transcardiac gradient (p=0.109). Similarly, subgroup analysis using a <30-day interval showed no significant association with an elevated transcoronary (p=0.13) or transcardiac gradient (p=0.15).

**Table 3A.** Mann-Whitney U Test PCAT FOR (RCA, LAD, LCx) as a predictor of detectable IL-1B.

| PCAT | U statistic | p-value | n detectable | n undetectable |
| --- | --- | --- | --- | --- |
| **RCA PCAT** | 78.0 | **0.323** | 5 | 25 |
| **LCx PCAT** | 63.0 | **0.889** | 5 | 25 |
| **LAD PCAT** | 67.0 | **0.716** | 5 | 25 |

**Table 3B.** Logistic regression for PCAT as a predictor of IL-1B. . Logistic regression modelling showed no significant relationship between PCAT attenuation and detectable IL-1β across any coronary vessel, including the RCA (OR 0.96, 95% CI 0.88–1.06, p = 0.456), LCx (OR 1.00, 95% CI 0.91–1.09, p = 0.941), and LAD (OR 0.99, 95% CI 0.90–1.08, p = 0.805) (Table 3B). Odds Ratios reflect the change in odds per 1 HU increase in PCAT HU.

| Predictor | OR | 95% CI | p-value |
| --- | --- | --- | --- |
| RCA PCAT | 0.96 | 0.88 – 1.06 | 0.456 |
| LCx PCAT | 1.00 | 0.91 – 1.09 | 0.941 |
| LAD PCAT | 0.99 | 0.90 – 1.08 | 0.805 |

## Discussion

This imaging/invasive study found no evidence of an association between coronary perivascular adipose tissue (PCAT) attenuation and intracardiac or intracoronary inflammatory cytokine gradients for either IL-6 or IL-1β (**Central illustration**). Across all coronary territories (RCA, LCx, LAD), PCAT attenuation did not predict transcardiac or transcoronary IL-6 gradients. Similarly, although IL-1β concentrations were frequently at or below the lower limit of detection, neither non-parametric group comparisons nor logistic regression examining the presence of detectable IL-1β demonstrated any relationship with PCAT attenuation. These findings remained consistent in subgroup analyses restricted to plaque-free vessels and across epicardial vessels, suggesting that the absence of association was not simply attributable to confounding by established atherosclerotic plaque. Taken together, these results suggest that CT-derived PCAT attenuation may not directly reflect contemporaneous soluble inflammatory cytokine gradients within the coronary circulation in this cohort. PCAT might capture inflammatory remodelling or structural perivascular changes that occur independently of immediate intracoronary cytokine gradients.

Notably, despite our cohort having a substantial burden of coronary artery disease with nearly half (44%) requiring revascularisation via PCI or CABG, we observed no robust relationship between PCAT attenuation and inflammatory cytokine gradients. This suggests that PCAT attenuation may represent a more complex or indirect marker of vascular risk, potentially reflecting chronic tissue remodelling, fibrosis, or longer-term structural changes rather than ongoing cytokine-mediated inflammation.

In this exploratory analysis, vessel-specific PCAT attenuation was not significantly associated with total plaque burden or individual plaque subtypes. Correlation coefficients were all non-significant suggesting that within this stable cohort, PCAT attenuation does not directly reflect quantitative plaque volume or compositional burden.

The plaque phenotype observed in this cohort was predominantly fibrotic, with relatively low non-calcified and fibrofatty components. This composition may potentially explain the absence of association between PCAT, and plaque subtypes typically linked to inflammatory activity. If PCAT reflects vascular inflammation rather than structural plaque burden per se, its relationship may be stronger in cohorts enriched for active lipid-rich, or high-risk plaque.

### Mechanistic link between PCAT and inflammation

The inflammatory hypothesis of coronary artery disease (CAD) is supported by previous trials showing targeted anti-inflammatory therapies to be associated with reductions in cardiovascular events independent of lipid lowering. The association of PCAT with inflammation is of significant clinical interest, because of its relation to plaque development and destabilisation.^21^ PCAT could identify phenotypes who are most likely to respond to new therapies in routine clinical practise^22^.

Contemporary literature increasingly supports PCAT its value as a prognostic and risk-stratification biomarker^1^, with multiple cohort studies have demonstrated that higher PCAT attenuation is associated with future myocardial infarction^4^, cardiac mortality, and adverse outcomes ^23^ even after adjustment for plaque burden and traditional risk factors. In this context - our findings add relevant nuance: while PCAT attenuation appears to carry cardiovascular risk prognostication, it may not primarily reflect active cytokine-driven plaque inflammation in patients with stable coronary disease.

While PCAT is recognised as a dynamic tissue that responds to inflammatory stimuli in an “inside-out” manner, particularly in response to acute coronary syndromes and myocardial infarction, there is also increasing interest also the “outside-in” hypothesis: whereby PCAT participates in atherogenic cascade; thus changes in PCAT radiodensity reflect coronary inflammatory biology in stable CAD through cytokine signalling, that promotes endothelial dysfunction, and plaque progression^24^.

Our results therefore challenge the assumption that elevated PCAT attenuation necessarily reflects active cytokine-driven coronary inflammation in stable disease. Instead, the biological signal captured by PCAT may not be mediated by sustained cytokine release. This interpretation is consistent with other negative mechanistic studies, including physiological investigations in INOCA cohorts^25^, where elevated PCAT attenuation has not been consistently associated with invasive coronary function abnormalities or higher rates of microvascular dysfunction.

### Importance of acuity

These findings do not invalidate PCAT attenuation as a prognostic marker of cardiovascular risk, but rather, they add insight into the contextual interpretation of PCAT. PCAT may not be uniformly metabolically active across all disease states, and its inflammatory phenotype may vary according to plaque biology and endothelial remodelling. It is possible that a threshold of extrinsic inflammatory stimulus is required before PCAT becomes biologically activated; however, our findings suggest that PCAT alone is unlikely to be the primary driver of inflammation, at least in stable disease states. Disentangling whether PCAT represents a driver of inflammation or a downstream marker is particularly challenging in the context of ACS, which is associated with elevated local cardiac cytokine production, including elevated IL-1β and IL-6 concentrations reported during acute events^26^. PCAT attenuation has also been shown to be greater around culprit lesions compared with stable segments^27^, raising the possibility that PCAT changes may reflect a reactive phenomenon in the setting of plaque destabilisation rather than baseline atherogenesis. The systemic inflammatory response accompanying myocardial injury further complicates interpretation of circulating biomarkers. For these reasons, we elected to study a stable outpatient cohort with recent CTCA (≤3 months), where acute ischaemic or injury-related inflammatory signals are less prominent, and where PCAT attenuation may more plausibly reflect chronic coronary biology and its potential role as an upstream contributor to local coronary inflammation.

### Limitations

While this study is the first to compare intracardiac and intracoronary cytokine sampling, which provides a more focused assessment of coronary biology than peripheral serum sampling alone, it has a number of limitations. First, the modest sample size limits power to detect small effect sizes. Cytokine assays, particularly for IL-1β, are constrained by lower limits of detection and biological variability. While a broader inflammatory panel encompassing additional cytokines and chemokines (e.g. TNF-α, MCP-1, IL-8, IL-10) may have provided further granularity, we opted for a more focused approach due limitations in blood volume sampling all sites, assay costs and sample processing complexity. Single time-point sampling may also fail to capture the dynamic nature of inflammatory signalling. It is also possible that inflammatory signalling between endothelium, and PCAT is highly localised and more paracrine, and there might not translate into measurable differences in cross-sectional cytokine concentrations within the coronary lumen or coronary sinus. Secondly, this stable CAD cohort demonstrated very low baseline inflammatory gradients (transcoronary: −0.04 ± 0.79 pg/mL; transcardiac: −0.01 ± 0.29 pg/mL). As the study deliberately excluded patients with acute coronary syndromes or unstable angina, this likely reflects a population with relatively low inflammatory activity. It is therefore plausible that measurable coronary cytokine gradients may only emerge in settings of acute plaque destabilisation, myocardial infarction, or active microvascular inflammation.

Future research should prioritise longitudinal evaluation to determine whether changes in PCAT attenuation over time reflect plaque evolution and whether these changes meaningfully modulate clinical outcomes. Understanding the biological context in which PCAT operates, including its interaction with plaque phenotype and inflammatory milieu - is necessary to refine its role beyond a broad risk marker. This will be particularly important in identifying the populations in whom PCAT has greatest clinical value, such as at-risk patients or lower risk individuals undergoing CTCA.

## Conclusion

The findings of this study demonstrate that coronary PCAT attenuation is not associated with transcardiac or transcoronary IL-6 or IL-1β gradients in patients with stable coronary artery disease, even in a cohort with significant anatomical disease burden and high rates of subsequent revascularisation. From a translational perspective these findings suggest that PCAT may represent a broader imaging signature of vascular vulnerability rather than a direct surrogate for real-time cytokine release. Further defining this distinction will be useful in delineating PCAT’s role as it is considered for incorporation into CT-based cardiovascular risk assessment frameworks.

## Data Availability

Available for serious collaboration on communication with the corresponding author

---

**Central Illustration.**
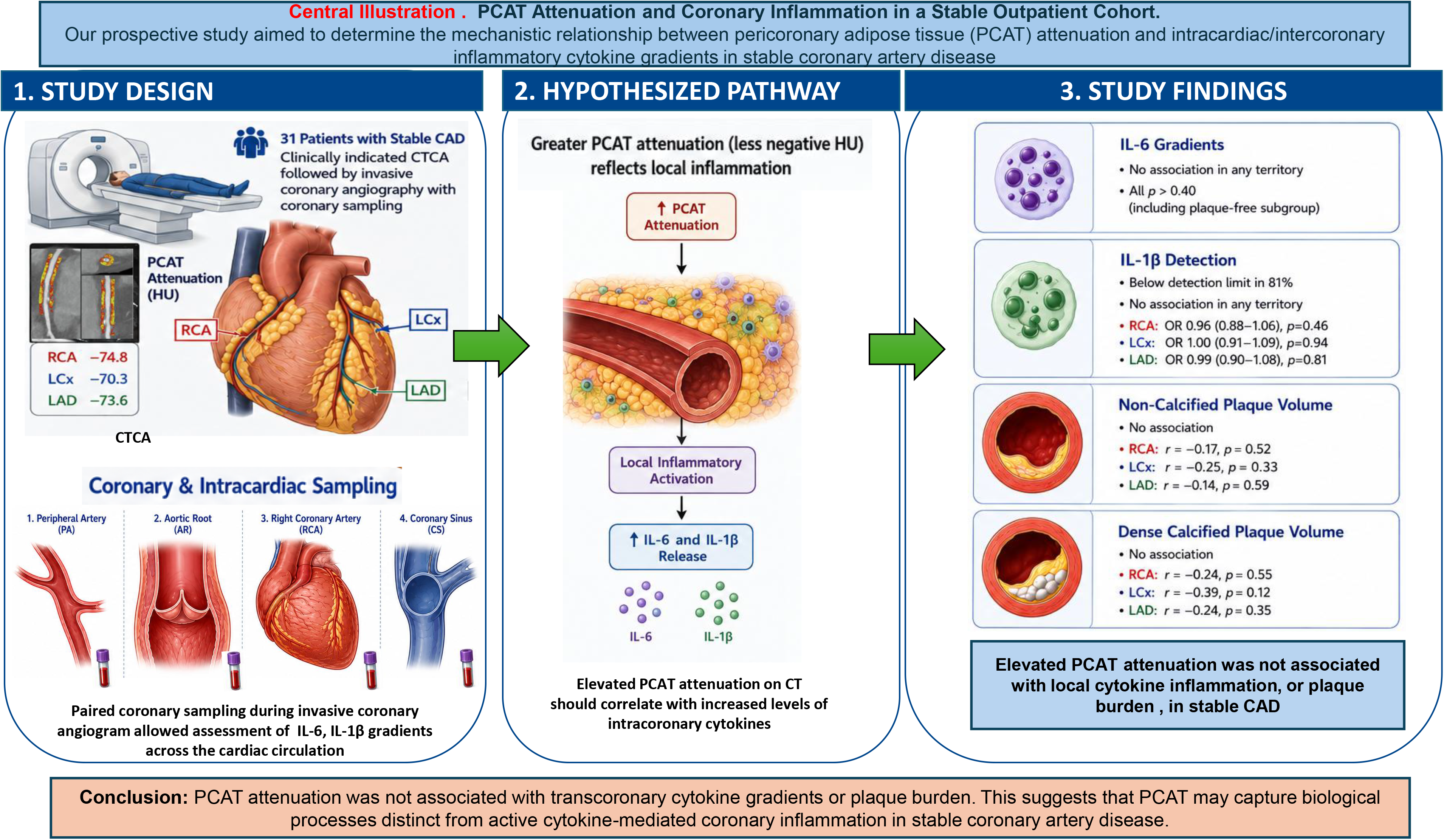
PCAT Attenuation and Coronary Inflammation in a Stable Outpatient Cohort. Our prospective study aimed to determine the mechanistic relationship between pericoronary adipose tissue (PCAT) attenuation and intracardiac/intercoronary inflammatory cytokine gradients in stable coronary artery disease

## Notes

### Competing Interest Statement

The authors have declared no competing interest.

### Author Declarations

Monash Human Research Ethics Committee

